# Different indicators of stress-induced hyperglycemia and poor outcomes after spontaneous intracerebral hemorrhage

**DOI:** 10.1101/2023.06.19.23291632

**Authors:** Xintong Song, Jia Zhang, Wenjuan Wang, Anxin Wang, Xiaoli Zhang, Sijia Li, Chuanying Wang, Xingquan Zhao, Qian Zhang

## Abstract

**Background:** This study aimed to compare the predictive value of metrics unique to stress-induced hyperglycemia (SIH) with fasting blood on poor functional outcomes of spontaneous intracerebral hemorrhage (sICH).

**Methods:** This investigation included 528 patients from a multicenter, observational, prospective cohort study. Poor functional outcomes were defined as modified Rankin Scale(mRS) ≥ 3. Logistic regression analyses were used to assess the relationship between indicators of SIH, including fasting blood glucose (FBG), two definitions of the stress hyperglycemia ratio [SHR, the fast blood glucose concentration/estimated average glucose (SHR1) and the ratio of glucose to HbA1c(SHR2)], and glycemic gap (GG), and poor functional outcomes at 30-day, 90-day, and 1-year.

**Results:** Higher values of all these indicators for SIH (e.g., FBG, GG, SHR1, and SHR2) were independently related to poor outcomes at 30-day, 90-day, and 1-year in patients with sICH (p < 0.05 for all models). Compared with the area under the curve (AUC), all these indicators performed greater AUC in predicting poor prognosis at 1-year (FBG: AUC=0.710; GG: AUC=0.741; SHR1: AUC=0.743) than 30-day and 90-day. And SHR2 has the highest predictive value among these indicators (AUC=0.748). Finally, diabetes had no statistical effect on the correlation between these indicators and poor functional outcomes at 30-day, 90-day, and 1-year in subgroup analysis (p for interaction >0.05).

**Conclusions:** Patients with higher FBG, GG, SHR1, and SHR2 values were more likely to have a poor functional outcome. SHR2 has the highest predictive value for poor outcomes at 30-day, 90-day and 1-year.

## Introduction

Spontaneous intracerebral hemorrhage (sICH), a subtype with significant morbidity and mortality, accounts for approximately 10% to 15% of all strokes^1^.Considering that the current effective treatment measures for sICH are limited, it is essential to find indicators that can predict the prognosis of patients with sICH and to some extent assist in the management of patients after hemorrhage^2^.

Hyperglycemia is commonly found in sICH patients on admission and previous studies have demonstrated that it increased mortality and poor functional outcomes in sICH patients^3, 4^.While hyperglycemia on admission may be caused by diabetes mellitus (DM) or stress state^5^. Stress-induced hyperglycemia(SIH) is transient hyperglycemia that occur after suffering an acute illness including stroke^6^.Previous studies have shown the relationship between SIH and increased occurrence of hemorrhagic transformation^7^,poor functional outcomes^8^,mortality^9^and infectious complications^10^in ICH patients.

However, SIH lacks a standardized definition and is a broad concept. It was usually defined according to blood glucose level in patients without previous diagnosed DM^6^.This definition has two shortcomings(1) It failed to distinguish between stress hyperglycemia and newly diagnosed or unidentified diabetes, and did not account for background glucose levels^11^. (2)Previous studies used different cutoff values of fasting blood glucose(FBG) ranging from 6.3 mmol/L to 8.6 mmol/L to define hyperglycemia ^4^. We need to find an indicator that corrects for chronic glycemic status to reflect the degree of SIH more accurately than FBG.

The stress hyperglycemia ratio (SHR) and the glycemic gap (GG) are two indicators which have been developed to assess SIH. Considering glycosylated hemoglobin (HbA1c) was a biochemical stability index representing about 2-3 months of mean glucose concentration^6^,SHR and GG plugged it into the calculation to remove the influence of chronic background glucose levels. And SHR has two definitions widely applied: the first one is the fasting blood glucose concentration/estimated average glucose (eAG)^12^,while the other one was the ratio of glucose to HbA1c^13^. GG is defined as the discrepancy between admission glucose level and eAG^14^. However, there is still no research on which indicator can more accurately predict functional outcomes of patients with sICH and become the most representative indicator of stress hyperglycemia in the future.

In this study, we prospectively explored the relationship between stress-induced hyperglycemia, assessed by FBG, GG, and the two definitions of SHR, and poor prognosis at 30 days, 90 days, and 1 year in patients with sICH. We also compared the predictive effect of these indicators, providing a basis for the selection of indicators in the subsequent research on stress hyperglycemia.

## Methods

### Study design and population

The current data for this study came from a multicenter, prospective, observational cohort study conducted at 13 hospitals in Beijing between January 2014 and September 2016. Data were collected by all participating centers and electronically delivered to the Coordination Center of Beijing Tiantan Hospital, Capital Medical University for combined analysis. This study was conducted by the guidelines from the Helsinki Declaration and was approved by the Institutional Review Board (IRB) of Beijing Tiantan Hospital, Capital Medical University. Every patent or their legal proxies provided a written informed consent.

The inclusion criteria of this cohort study were (1) ICH was diagnosed based on the World Health Organization standard and confirmed by CT scan, (2) first-ever acute-onset ICH, (3) age≥18 years old and (4) arriving at hospital within 72h after onset^15^. After excluding patients with major comorbidities or late-stage diseases, 1964 eligible patients were included into this cohort. In this research, we further exclude patients with the following diagnoses(1) primary ventricular hemorrhage; (2) secondary ICH, which caused by cerebrovascular malformations, trauma, tumors, aneurysms, cerebral venous thrombosis, congenital or acquired coagulation disorders, or hemorrhagic transformation after ischemic stroke; and reject patients (3) whose fasting blood glucose and HbA1c information were not available; (4) without complete relevant covariate data(e.g., hypertension, diabetes mellitus, history of cerebral infarction, history of myocardial infarction, mRS score before onset, GCS score, NIHSS scores, location of hematoma, hematoma volume, and whether breaking into ventricle, or (5) without follow-up information. 528 patients were eventually included in this study (**Figure 1**).

**Figure 1.**
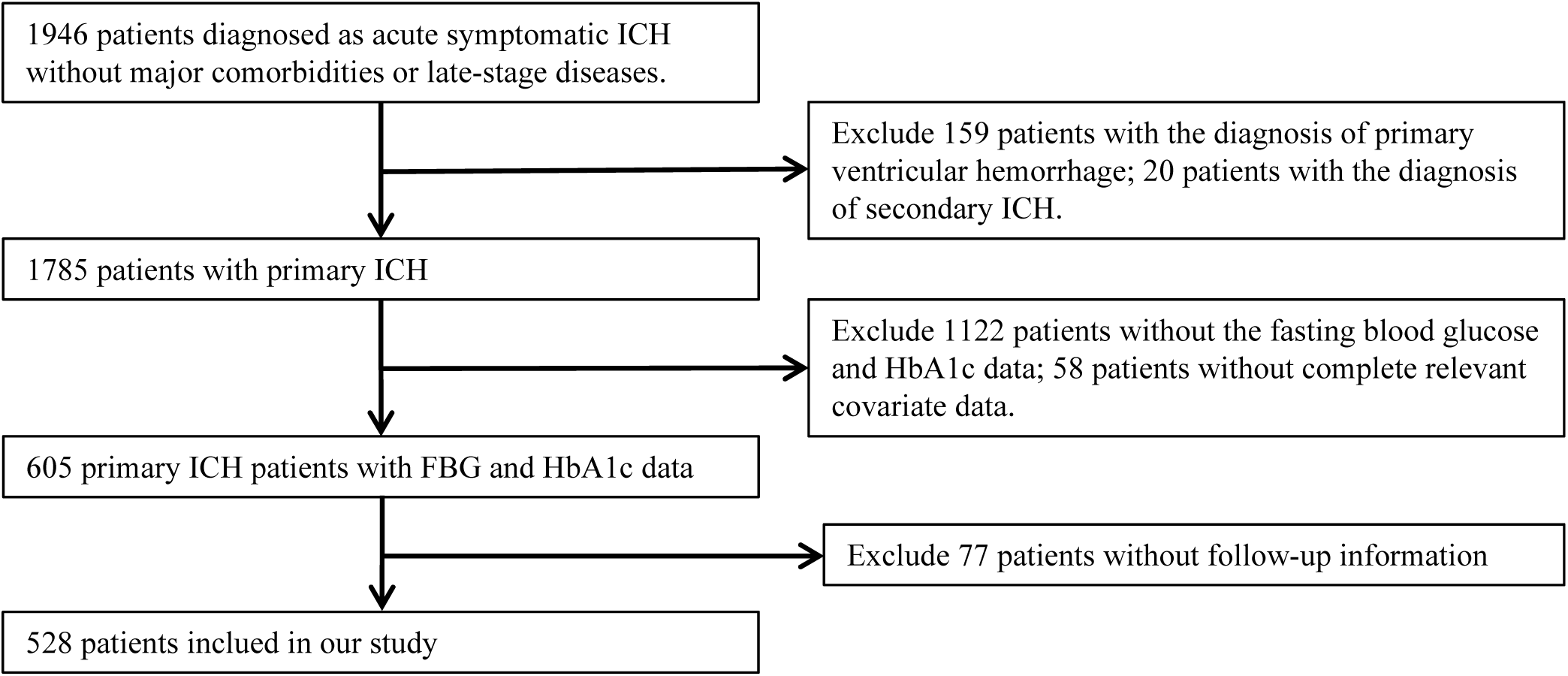
flowchart of study patients.

### Clinical data collection

Using standard questionnaires, the clinical data of patients concerning demographics characteristics (e.g., age and sex), health habits (e.g., alcohol consumption and smoking), medical history (e.g., hypertension, diabetes mellitus, dyslipidemia, history of myocardial infarction and cerebral infarction,) and prior medication use(antihypertensive, antidiabetic, lipid-lowering, antiplatelet, and anticoagulant)were documented. The modified Rankin Scale (mRS) scores before the onset of ICH, admission National Institutes of Health Stroke Scale (NIHSS) score, and admission Glasgow Coma Scale (GCS) score were all collected by qualified physicians. The medical system was applied for figuring out the diagnosis at discharge. The ICH Score, which is plotted to evaluate ICH severity, was calculated as the sum of individual points allocated to GCS score, age, infratentorial origin, ICH volume, and intraventricular hemorrhage (IVH) ^16^.Possessing an identifiable history or with diagnosis at discharge was considered to have hypertension and dyslipidemia. Diabetes referred to having a self-reported history, admission HbA1c ≥ 6.5%, or with a discharge diagnosis of diabetes^15^.

### Laboratory tests and the evaluation of stress hyperglycemia

All laboratory indicators (including FBG and Hb1Ac) was assessed the following morning after an overnight fasting for at least 8 hours. Other laboratory tests, including creatinine, total cholesterol (TC), high-density lipoprotein cholesterol (HDL-C), low-density lipoprotein cholesterol (LDL-C), alanine aminotransferase(ALT), systolic blood pressure(SBP), and diastolic blood pressure(DBP) were also evaluated during admission. Estimated average blood glucose level using the equation eAG =28.7 * HbA1c–46.7 reflects glucose control from the previous 3 months^17^. Stress-induced hyperglycemia was evaluated by SHR and GG calculating using the following formulas: SHR1=FBG(mg/dl)/eAG, SHR2=FBG(mg/dl)/ HbA1c (%), and GG=FBG(mg/dl)-eAG^14^.

### Neuroimaging

To determine hematoma characteristics, first computerized tomography (CT) scans were finished in 24 hours after admission. Two investigators were blinded to analyze imaging data separately. The ABC/2 method was applied to calculate the hemorrhage volume and the calculation process has been described previously^18, 19^.

### Follow-up information and clinical outcomes

Well-trained investigators performed regular telephone calls to assess the scores of mRS at 30 days, 90 days, and 1 year after sICH onset. Clinical information of patients was hidden from all research personals. The definition of poor functional outcomes was mRS ≥ 3.

### Statistics analysis

Statistical analyses were completed with SAS software version 9.4 (SAS Institute), and the statistical tests in our study were two-sided with p < 0.05 for statistical significance. First, patients were divided into two groups based on their functional outcome at 30 days, 90 days, and 1 year. Data for continuous variables were described as means ± standard deviation (SD), while categorical variables were expressed as numbers (proportions). Student’s t-test or Mann-Whitney U-test used for continuous variables and chi-squared test for categorical variables when comparing the differences among the two groups. Then, we identified the clinical confounding risk factors that were documented to be related with poor functional outcomes in sICH patients and possible confounders with p<0.05 in analysis of baseline characteristics between two groups^20^.Logistic regression model was established taking above confounders into consideration to generate odds ratios (OR) with 95% confidence intervals(CI) for the link between indicators (SHR1, SHR2, and GG) and poor clinical outcomes. The crude model was univariable analysis. Age and sex were adjusted in model 1, and model 2 was adjusted for age, sex, hypertension, dyslipidemia, diabetes, history of cerebral infarction, history of myocardial infarction, prior anticoagulant agents, prior antiplatelet use, prior antihypertensive agents, and mRS before onset. In model 3, we additional adjustments was made for admission NIHSS score, admission GCS score, systolic blood pressure, admission ALT, admission HDL-C and admission LDL-C. Model 4 was adjusted for covariates in model 3 and hematoma characteristics such as presence of intraventricular extension, location of hematoma, and hematoma volume. What’s more, subgroup analyses were performed to investigate the relationship between SIH and poor functional outcomes at different follow-up periods in patient with or without DM. Additionally Receiver operating characteristic (ROC) curves were made to assess predictive values of these indicators for poor clinical outcomes at 30 days, 90 days, and 1 year separately. Area under the curve (AUC) was used to evaluate accuracy of prediction. Finally, C-statistic, net reclassification improvement (NRI) index and integrated discrimination improvement (IDI) index were conducted to analyses the incremental predictive value of these indicators beyond the ICH score^16^.

## Results

### Baseline characteristics

Among 528 sICH patients, all were successfully followed up with 30 days and 90 days, 495 of whom were still in contact at 1 year. Therefore, we limited 1-year analysis to 495 patients and performed majority of our analyses on 528 patients. This analysis included 528 sICH patients (374 males and 154 females) with the average age of 59.02±13.04 years. There were 280(53.03%), 232(43.94%) and 188(37.98%) patients with poor functional outcomes(mRS≥3) at 30 days, 90 days and 1 year respectively.

Patients with poor functional outcomes were statistically older(p<0.001), had lower proportion of dyslipidemia(p<0.05), higher proportion of prior anticoagulant use history(p<0.05), higher mRS score before onset(p<0.05) than patents with good clinical outcomes. And they also had higher admission SBP(p<0.05), higher HDL-C(p<0.05), lower admission GCS score(p<0.001), higher admission NHISS score(p<0.001), higher hematoma volume(p<0.001), and a larger proportion of intraventricular extension(p<0.001). At 90-day and 1-year follow-up, people with poor prognosis had higher proportion of previous antiplatelet(p<0.05), antihypertensive(p<0.05), and cerebral infarction history(p<0.05), but the difference was not significant at 30-day follow-up. **Table 1** lists the clinical characteristics of patients in this analysis.

**Table 1.**
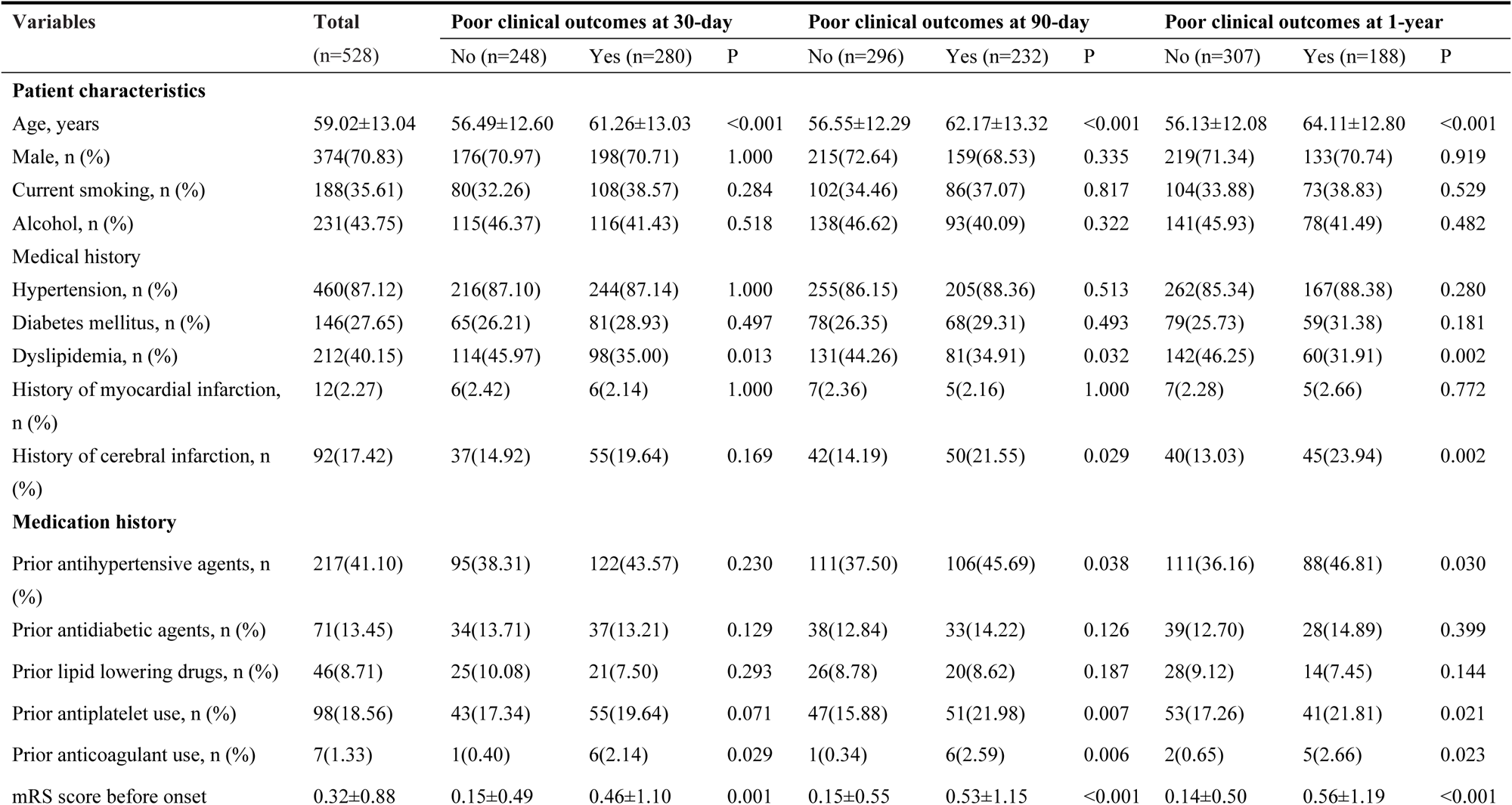
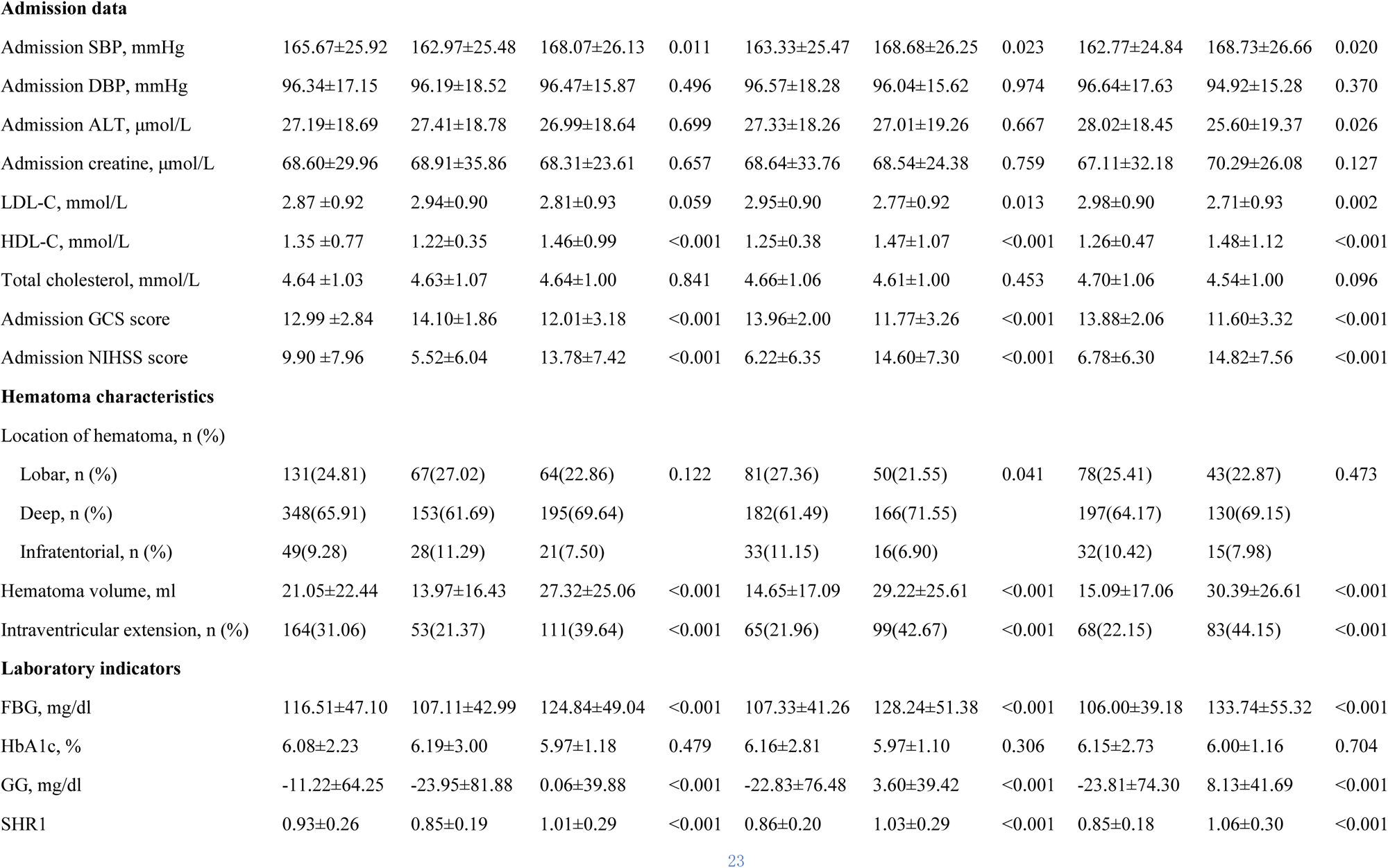
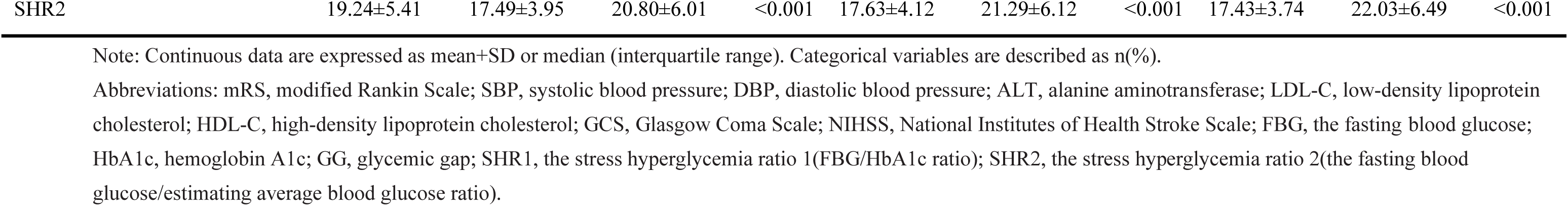
Baseline characteristics.

| Variables | Total<br>(n=528) | Poor clinical outcomes at 30-day |  |  | Poor clinical outcomes at 90-day |  |  | Poor clinical outcomes at 1-year |  |  |
| --- | --- | --- | --- | --- | --- | --- | --- | --- | --- | --- |
|  |  | No (n=248) | Yes (n=280) | P | No (n=296) | Yes (n=232) | P | No (n=307) | Yes (n=188) | P |
| Patient characteristics |  |  |  |  |  |  |  |  |  |  |
| Age, years | 59.02±13.04 | 56.49±12.60 | 61.26±13.03 | <0.001 | 56.55±12.29 | 62.17±13.32 | <0.001 | 56.13±12.08 | 64.11±12.80 | <0.001 |
| Male, n (%) | 374(70.83) | 176(70.97) | 198(70.71) | 1.000 | 215(72.64) | 159(68.53) | 0.335 | 219(71.34) | 133(70.74) | 0.919 |
| Current smoking, n (%) | 188(35.61) | 80(32.26) | 108(38.57) | 0.284 | 102(34.46) | 86(37.07) | 0.817 | 104(33.88) | 73(38.83) | 0.529 |
| Alcohol, n (%) | 231(43.75) | 115(46.37) | 116(41.43) | 0.518 | 138(46.62) | 93(40.09) | 0.322 | 141(45.93) | 78(41.49) | 0.482 |
| Medical history |  |  |  |  |  |  |  |  |  |  |
| Hypertension, n (%) | 460(87.12) | 216(87.10) | 244(87.14) | 1.000 | 255(86.15) | 205(88.36) | 0.513 | 262(85.34) | 167(88.38) | 0.280 |
| Diabetes mellitus, n (%) | 146(27.65) | 65(26.21) | 81(28.93) | 0.497 | 78(26.35) | 68(29.31) | 0.493 | 79(25.73) | 59(31.38) | 0.181 |
| Dyslipidemia, n (%) | 212(40.15) | 114(45.97) | 98(35.00) | 0.013 | 131(44.26) | 81(34.91) | 0.032 | 142(46.25) | 60(31.91) | 0.002 |
| History of myocardial infarction, n (%) | 12(2.27) | 6(2.42) | 6(2.14) | 1.000 | 7(2.36) | 5(2.16) | 1.000 | 7(2.28) | 5(2.66) | 0.772 |
| History of cerebral infarction, n (%) | 92(17.42) | 37(14.92) | 55(19.64) | 0.169 | 42(14.19) | 50(21.55) | 0.029 | 40(13.03) | 45(23.94) | 0.002 |
| Medication history |  |  |  |  |  |  |  |  |  |  |
| Prior antihypertensive agents, n (%) | 217(41.10) | 95(38.31) | 122(43.57) | 0.230 | 111(37.50) | 106(45.69) | 0.038 | 111(36.16) | 88(46.81) | 0.030 |
| Prior antidiabetic agents, n (%) | 71(13.45) | 34(13.71) | 37(13.21) | 0.129 | 38(12.84) | 33(14.22) | 0.126 | 39(12.70) | 28(14.89) | 0.399 |
| Prior lipid lowering drugs, n (%) | 46(8.71) | 25(10.08) | 21(7.50) | 0.293 | 26(8.78) | 20(8.62) | 0.187 | 28(9.12) | 14(7.45) | 0.144 |
| Prior antiplatelet use, n (%) | 98(18.56) | 43(17.34) | 55(19.64) | 0.071 | 47(15.88) | 51(21.98) | 0.007 | 53(17.26) | 41(21.81) | 0.021 |
| Prior anticoagulant use, n (%) | 7(1.33) | 1(0.40) | 6(2.14) | 0.029 | 1(0.34) | 6(2.59) | 0.006 | 2(0.65) | 5(2.66) | 0.023 |
| mRS score before onset | 0.32±0.88 | 0.15±0.49 | 0.46±1.10 | 0.001 | 0.15±0.55 | 0.53±1.15 | <0.001 | 0.14±0.50 | 0.56±1.19 | <0.001 |
| **Admission data** |  |  |  |  |  |  |  |  |  |  |
| Admission SBP, mmHg | 165.67±25.92 | 162.97±25.48 | 168.07±26.13 | 0.011 | 163.33±25.47 | 168.68±26.25 | 0.023 | 162.77±24.84 | 168.73±26.66 | 0.020 |
| Admission DBP, mmHg | 96.34±17.15 | 96.19±18.52 | 96.47±15.87 | 0.496 | 96.57±18.28 | 96.04±15.62 | 0.974 | 96.64±17.63 | 94.92±15.28 | 0.370 |
| Admission ALT, μmol/L | 27.19±18.69 | 27.41±18.78 | 26.99±18.64 | 0.699 | 27.33±18.26 | 27.01±19.26 | 0.667 | 28.02±18.45 | 25.60±19.37 | 0.026 |
| Admission creatine, μmol/L | 68.60±29.96 | 68.91±35.86 | 68.31±23.61 | 0.657 | 68.64±33.76 | 68.54±24.38 | 0.759 | 67.11±32.18 | 70.29±26.08 | 0.127 |
| LDL-C, mmol/L | 2.87 ±0.92 | 2.94±0.90 | 2.81±0.93 | 0.059 | 2.95±0.90 | 2.77±0.92 | 0.013 | 2.98±0.90 | 2.71±0.93 | 0.002 |
| HDL-C, mmol/L | 1.35 ±0.77 | 1.22±0.35 | 1.46±0.99 | <0.001 | 1.25±0.38 | 1.47±1.07 | <0.001 | 1.26±0.47 | 1.48±1.12 | <0.001 |
| Total cholesterol, mmol/L | 4.64 ±1.03 | 4.63±1.07 | 4.64±1.00 | 0.841 | 4.66±1.06 | 4.61±1.00 | 0.453 | 4.70±1.06 | 4.54±1.00 | 0.096 |
| Admission GCS score | 12.99 ±2.84 | 14.10±1.86 | 12.01±3.18 | <0.001 | 13.96±2.00 | 11.77±3.26 | <0.001 | 13.88±2.06 | 11.60±3.32 | <0.001 |
| Admission NIHSS score | 9.90 ±7.96 | 5.52±6.04 | 13.78±7.42 | <0.001 | 6.22±6.35 | 14.60±7.30 | <0.001 | 6.78±6.30 | 14.82±7.56 | <0.001 |
| Lobar, n (%) | 131(24.81) | 67(27.02) | 64(22.86) | 0.122 | 81(27.36) | 50(21.55) | 0.041 | 78(25.41) | 43(22.87) | 0.473 |
| Deep, n (%) | 348(65.91) | 153(61.69) | 195(69.64) |  | 182(61.49) | 166(71.55) |  | 197(64.17) | 130(69.15) |  |
| Infratentorial, n (%) | 49(9.28) | 28(11.29) | 21(7.50) |  | 33(11.15) | 16(6.90) |  | 32(10.42) | 15(7.98) |  |
| Hematoma volume, ml | 21.05±22.44 | 13.97±16.43 | 27.32±25.06 | <0.001 | 14.65±17.09 | 29.22±25.61 | <0.001 | 15.09±17.06 | 30.39±26.61 | <0.001 |
| Intraventricular extension, n (%) | 164(31.06) | 53(21.37) | 111(39.64) | <0.001 | 65(21.96) | 99(42.67) | <0.001 | 68(22.15) | 83(44.15) | <0.001 |
| **Laboratory indicators** |  |  |  |  |  |  |  |  |  |  |
| FBG, mg/dl | 116.51±47.10 | 107.11±42.99 | 124.84±49.04 | <0.001 | 107.33±41.26 | 128.24±51.38 | <0.001 | 106.00±39.18 | 133.74±55.32 | <0.001 |
| HbA1c, % | 6.08±2.23 | 6.19±3.00 | 5.97±1.18 | 0.479 | 6.16±2.81 | 5.97±1.10 | 0.306 | 6.15±2.73 | 6.00±1.16 | 0.704 |
| GG, mg/dl | -11.22±64.25 | -23.95±81.88 | 0.06±39.88 | <0.001 | -22.83±76.48 | 3.60±39.42 | <0.001 | -23.81±74.30 | 8.13±41.69 | <0.001 |
| SHR1 | 0.93±0.26 | 0.85±0.19 | 1.01±0.29 | <0.001 | 0.86±0.20 | 1.03±0.29 | <0.001 | 0.85±0.18 | 1.06±0.30 | <0.001 |
| SHR2 | 19.24±5.41 | 17.49±3.95 | 20.80±6.01 | <0.001 | 17.63±4.12 | 21.29±6.12 | <0.001 | 17.43±3.74 | 22.03±6.49 | <0.001 |

Note: Continuous data are expressed as mean+SD or median (interquartile range). Categorical variables are described as n(%).
Abbreviations: mRS, modified Rankin Scale; SBP, systolic blood pressure; DBP, diastolic blood pressure; ALT, alanine aminotransferase; LDL-C, low-density lipoprotein cholesterol; HDL-C, high-density lipoprotein cholesterol; GCS, Glasgow Coma Scale; NIHSS, National Institutes of Health Stroke Scale; FBG, the fasting blood glucose; HbA1c, hemoglobin A1c; GG, glycemic gap; SHR1, the stress hyperglycemia ratio 1(FBG/HbA1c ratio); SHR2, the stress hyperglycemia ratio 2(the fasting blood glucose/estimating average blood glucose ratio).

### The correlation between SIH indicators and the poor clinical outcomes

It was presented that sICH patients with poor outcomes had much higher FBG, (107.11±42.99 vs. 124.84±49.04, *p*<0.001), higher GG (-23.95±81.88 vs. 0.06±39.88, p<0.001), higher SHR1(0.85±0.19 vs. 1.01±0.29, p<0.001) and higher SHR2 (17.49±3.95 vs. 20.80±6.01, p<0.001) comparing with those with good outcomes at 30 days. While there was no significant difference in blood HBA1c level between poor and good outcomes (6.19±3.00 vs. 5.97±1.18, p=0.479) at 30 days, which is another important indicator of glucose metabolism. Similar outcomes were observed at 90 days and 1 year follow-up, which showed in **Table 1**.

Univariate logistic regression analysis suggested that higher level of all the indicators in our current study, including FBG(Q3 vs. Q1: OR = 3.866, 95% CI [2.486,6.011],p<0.0001),GG(Q3 vs. Q1: OR = 4.554, 95% CI [2.900,7.152],p<0.0001), SHR1(Q3 vs. Q1: OR = 5.027, 95% CI [3.192,7.919],p<0.0001), and SHR2(Q3 vs. Q1: OR = 4.520, 95% CI [2.883,7.087],p<0.0001) were significantly related to the risk of poor clinical outcomes at 30 days. (**Table 2**) The association between these indicators and poor clinical outcomes still existed (p<0.05) after adjusting for confounding risk factors. Similar results were observed in 90-day and 1-year outcomes (all p < 0.05). Additionally, it also showed a significant increasing trend of FBG, GG, SHR1 and SHR2 in our analysis (*p* for trend < 0.01 for all models). (**Supplementary Table S1 - 3.**)

**Table 2.** Univariate and multivariate logistic regression analyses for 30-day, 90-day, and 1-year poor functional outcomes.

| Variables |  | 30-day poor functional outcomes |  | 90-day poor functional outcomes |  | 1-year poor functional outcomes |  |
| --- | --- | --- | --- | --- | --- | --- | --- |
|  |  | Unadjusted OR<br>(95% CI) | Adjusted OR<br>(95% CI) | Unadjusted OR<br>(95% CI) | Adjusted OR<br>(95% CI) | Unadjusted OR<br>(95% CI) | Adjusted OR<br>(95% CI) |
| FBG, mg/dl | Q1($\leq 93.78$) | reference | reference | reference | reference | reference | reference |
|  | Q2(94.32-116.46) | 2.701(1.752,4.165) | 2.530(1.402,4.567) | 2.396(1.526,3.761) | 1.970(1.057,3.672) | 2.507(1.515,4.150) | 2.145(1.033,4.454) |
| | Q3($\geq 117.00$) | 3.866(2.486,6.011) | 2.602(1.203,5.629) | 4.557(2.894,7.173) | 3.566(1.625,7.827) | 6.562(3.983,10.812) | 8.582(3.554,20.724) |
|  | P for trend | <0.0001 | 0.0053 | <0.0001 | 0.0013 | <0.0001 | <0.0001 |
| SHR1 | Q1($\leq 0.80$) | reference | reference | reference | reference | reference | reference |
|  | Q2(0.80-0.99) | 2.018(1.315,3.099) | 1.568(0.864,2.845) | 2.176(1.382,3.427) | 1.542(0.816,2.915) | 2.283(1.373,3.798) | 1.644(0.787,3.433) |
| | Q3($\geq 0.99$) | 5.027(3.192,7.919) | 3.379(1.726,6.616) | 5.516(3.480,8.742) | 3.206(1.641,6.262) | 8.031(4.823,13.372) | 6.606(3.064,14.243) |
|  | P for trend | <0.0001 | 0.0004 | <0.0001 | 0.0006 | <0.0001 | <0.0001 |
| SHR2 | Q1($\leq 16.55$) | reference | reference | reference | reference | reference | reference |
|  | Q2(16.57-20.09) | 1.920(1.253,2.943) | 1.452(0.798,2.641) | 2.176(1.382,3.427) | 1.531(0.808,2.903) | 2.315(1.382,3.877) | 1.628(0.770,3.440) |
| | Q3($\geq 20.11$) | 4.520(2.883,7.087) | 3.018(1.541,5.912) | 5.516(3.480,8.742) | 3.480(1.759,6.884) | 8.614(5.158,14.388) | 7.446(3.428,16.175) |
|  | P for trend | <0.0001 | 0.0014 | <0.0001 | 0.0003 | <0.0001 | <0.0001 |
| GG, mg/dl | Q1($\leq -24.03$) | reference | reference | reference | reference | reference | reference |
|  | Q2(-24.00- -1.12) | 1.669(1.091,2.555) | 1.321(0.726,2.404) | 1.761(1.125,2.758) | 1.372(0.725,2.597) | 1.971(1.193,3.255) | 1.706(0.816,3.569) |
| | Q3($\geq -1.06$) | 4.554(2.900,7.152) | 3.344(1.707,6.553) | 4.903(3.115,7.717) | 3.218(1.645,6.296) | 7.238(4.376,11.971) | 6.733(3.124,14.511) |
|  | P for trend | <0.0001 | 0.0005 | <0.0001 | 0.0006 | <0.0001 | <0.0001 |

Multivariate logistic analysis was adjusted for age, sex, hypertension, diabetes, dyslipidemia, history of cerebral infarction, history of myocardial infarction, prior anticoagulant agents, prior antihypertensive agents, prior antiplatelet use, mRS before onset, admission NIHSS score, admission GCS score, SBP, admission ALT, HDL-C, LDL-C, location of hematoma, hematoma volume and intraventricular extension.
FBG, the fasting blood glucose; SHR1, the stress hyperglycemia ratio 1(the fasting blood glucose /HbA1c ratio);SHR2, the stress hyperglycemia ratio 2(the fasting blood glucose/estimating average blood glucose ratio); GG, glycemic gap; HbA1c, hemoglobin A1c; mRS, modified Rankin Scale; NIHSS, National Institutes of Health Stroke Scale; GCS, Glasgow Coma Scale; ALT, alanine aminotransferase; LDL-C, low-density lipoprotein cholesterol; HDL-C, high-density lipoprotein cholesterol; SBP, systolic blood pressure.

### Predictive value of different indicators for poor functional outcomes

ROC curves of independent predictors of poor outcomes at different periods after sICH onset were drawn, and the AUCs, 95% CIs, and *p*-values of different indicators were shown in **Table 3**. By comparing the AUCs of these several indexes to show the accuracy of their predictive effects, we found that SHR2 had the highest AUC for adverse prognostic outcomes at 30-day (AUC=0.685,95%CI:0.640-0.730, p<0.0001), 90-day (AUC=0.705,95%CI:0.661-0.750,p<0.0001), and 1-year (AUC=0.748,95%CI:0.702-0.794, p<0.0001) after the onset of sICH. And all of these indicators performed greatest AUC in predicting poor prognosis at 1-year (FBG: AUC=0.710, 95%CI:0.663-0.757; GG: AUC=0.741, 95%CI:0.694-0.787; SHR1: AUC=0.743, 95%CI:0.697-0.789).

**Table 3.** Receiver operating characteristic analysis of different indicators for the prediction of poor functional outcome at 30-day,90-day and 1-year.

| Poor functional outcomes | Variables | Youden | Cutoff value | AUC (95%CI) | p-value |
| --- | --- | --- | --- | --- | --- |
| 30-day | FBG, mg/dl | 0.311 | 99.54 | 0.666(0.619,0.712) | <0.0001 |
|  | GG, mg/dl | 0.300 | -4.77 | 0.677(0.631,0.722) | <0.0001 |
|  | SHR1 | 0.300 | 0.96 | 0.681(0.635,0.726) | <0.0001 |
|  | SHR2 | 0.306 | 18.53 | 0.685(0.640,0.730) | <0.0001 |
| 90-day | FBG, mg/dl | 0.301 | 99.54 | 0.675(0.630,0.721) | <0.0001 |
|  | GG, mg/dl | 0.321 | -12.11 | 0.697(0.651,0.743) | <0.0001 |
|  | SHR1 | 0.318 | 0.94 | 0.700(0.655,0.745) | <0.0001 |
|  | SHR2 | 0.326 | 19.32 | 0.705(0.661,0.750) | <0.0001 |
| 1-year | FBG, mg/dl | 0.364 | 102.78 | 0.710(0.663,0.757) | <0.0001 |
|  | GG, mg/dl | 0.397 | -6.80 | 0.741(0.694,0.787) | <0.0001 |
|  | SHR1 | 0.403 | 0.94 | 0.743(0.697,0.789) | <0.0001 |
|  | SHR2 | 0.417 | 19.08 | 0.748(0.702,0.794) | <0.0001 |

AUC, area under the curve; CI, confidence intervals; FBG, the fasting blood glucose; SHR1, the stress hyperglycemia ratio 1(the fasting blood glucose /HbA1c ratio); SHR2, the stress hyperglycemia ratio 2(the fasting blood glucose/estimating average blood glucose ratio); GG, glycemic gap; HbA1c, hemoglobin A1c.

The cutoff values of SHR2(18.53 at 30-day, 19.32 at 90-day, 19.08 at 1-year) and GG (-4.7 mg/dl at 30-day, -12.11 mg/dl at 90-day and -6.8 mg/dl at 1-year) were inconsistent at different follow-up time. While FBG had same cutoff value (99.54 mg/dl) at 30-day and 90-day, and the cutoff value of SHR1 at 90-day and 1-year was 0.94.

### Results of subgroup analysis and incremental predictive value

In a subgroup analysis of patients with or without diabetes, the results suggested that diabetes didn’t influence the association between these indicators (FBG, GG, SHR1, and SHR2) and poor clinical outcomes at 30-day (p for interaction>0.05). Although 64 observations with diabetes and 26 observations without diabetes were deleted due to missing values for the response or explanatory variables, making some invalid data in subgroup analysis for 1-year, this phenomenon was also seen in the prediction of outcomes at 90-day and 1-year follow-up (p for interaction >0.05) in **Supplementary Table S4**.

The incremental value of FBG, SHR1, SHR2 and GG for predicting poor functional outcomes at 30-day,90-day and 1-year was showed in **Supplementary Table S5**. When GG, SHR1, SHR2 and FBG were added to ICH scores, the predictive effect of poor prognosis was significantly increased in C-statistics, NRI statistics, and IDI statistics.

## Discussion

We found that patients with higher FBG, GG, SHR1, and SHR2 values were more likely to experience poor functional outcomes at 30 days, 90 days, and 1 year period. All indicators we studied performed best in predicting 1-year adverse prognosis, and SHR2 had strongest predictive value for 30-day,90-day, and 1-year poor functional outcomes among the four indicators. Additionally, the association between all these indicators and poor functional results at 30-day,90-day, and 1-year was unaffected by diabetes, as we unexpectedly found in subgroup analysis.

Previous studies suggested hyperglycemia was associated with brain edema and neuronal apoptosis^21, 22^, hematoma expansion^23^, short-term and long-term mortality^24, 25^, and poor functional outcomes^3, 5, 22, 24, 26^ in ICH patients. Hyperglycemia in a hospitalized patient may be a sign of poorly controlled chronic diabetes, a transient physiologic response to a concurrent illness (stress hyperglycemia) or be a combination of both^12^. SIH, a greater alternation caused by sICH, may play as a better predictor when comparing with background status like diabetes. So, more and more work has begun to explore the relationship between SIH and spontaneous cerebral hemorrhage.

Many metrics have been considered as an indicator representing the extent of SIH. Conventionally, SIH can be identified by FBG ≥10 mmol/L in hospitalized non-diabetic patients, which is the recommended threshold for hyperglycemic treatment^27^.As a marker of SIH, FBG can partially reflect the changes in blood glucose, but it cannot effectively be employed as a diagnostic of SIH in a patient cohort with a wide range of HbA1c and a large variation in background blood glucose^28^. To better quantify stress hyperglycemia, SHR was first proposed by Roberts et al^12^used HbA1c to calculate background glycemia, which is calculate by FBG /eAG. It has been indicated that SHR1 is a credible predictor for early hematoma expansion and poor outcomes in previous study^8^. However, due to controversy persisting over eAG as a systematically biased estimate of self-monitored mean blood glucose, some scholars have noted that eAG should be utilized cautiously in clinical practice^29^. A more applicable and prevalent definition of SHR(SHR2) was the ratio of glucose to HbA1c ^30^. Furthermore, Li et al.^31^ revealed that SHR2 was independently related to poor functional outcomes at discharge and 3 months in patients with ICH. Glycemic gap (GG) as another indicator for evaluating SIH is defined as admission blood glucose – eAG. Previous study showed that GG was a good discriminative power in predicting in-hospital mortality in diabetes patients with ICH^9^. The above four glucose metrics, including FBG, GG, SHR1 and SHR2, widely accepted as SIH indicators, can represent acute changes in blood glucose levels caused by SIH when taking premorbid background glycemia into account. We demonstrated that they were all significantly correlated with poor functional outcomes of sICH at 30 days, 90 days, and 1 year, and can all be useful predictive indicators of poor long-term and short-term outcomes of sICH in our analysis.

However, there is a lack of comparison on the predictive effects of the four markers. In our study we innovatively solved this problem and reached the following conclusion: the SHR2 exhibited a stronger predictive value for poor clinical outcomes of sICH at 30-day, 90-day, and 1-year compared with the other three. It suggested that SHR2 could more precisely reflect physiological stress and serve as a better predictive indicator in sICH patients compared to FBG, GG and SHR1.

Different literatures currently used inconsistent cut-off to define SIH. Sensitivity tests around these thresholds are incomplete, and some of them didn’t provide validation of improved prognostic insight. The FBG values used to define SIH ranged from 140 mg/dL (7.8 mmol/L) to 200 mg/dL (11.1 mmol/L)^32–34^. One of the most cited definitions is set as a numerical value, cut-off >140 mg/dL (7.77 mmol/L)^35^. We also applied ROC curves to find optimal SIH cut-offs for patients with poor functional outcomes at 30-day(99.45mg/dl), 90-day (99.45 mg/dl) and 1-year (102.78 mg/dl), which is not consistent with past standards. For GG, one cohort for sICH patients used a threshold of 30.0 mg/dl to define SIH that predicted mortality with area under the curve (AUC=0.655)^36^. While our study found that -4.77 mg/dl was the ROC curve-derived GG cut-off value for poor outcomes at 30 days, while it was - 12.11 mg/dl 90 days and-6.8 mg/dl at 1 year. Moreover, Ye et.al used ROC analysis to estimate the ability of SHR1 to predict the remote diffusion-weighted imaging lesion (R-DWILs) occurrence and got 0.83 as cutoff value of SHR1^37^. The cut-off points of SHR1 for ICH patients for predicting secondary neurological deterioration within 48h, 30-day mortality, and 3-month poor functional outcomes were 1.067, 1.226, and 0.905 in another study^8^. And the cutoff value we obtained for poor prognosis of sICH population is between the two studies, 0.96 at 30-day,0.94 at 90-day and 1-year, respectively. Li et.al chose the median of the glucose-to-HbA1c ratio (SHR2) as a threshold of which was 1.02 and found higher SHR2 (≥1.02) remained to be independently correlated with poor functional outcomes at discharge and 90 days ^31^. Our study got 18.53,19.32 and 19.08 as cutoff value for poor clinical outcomes at 30 days,90 days and 1year, respectively. Because there is no unified standard for cutoff values as threshold for SIH and the cutoff values we studied were not consistent in different follow-up periods, more studies should be concerned to propose the appropriate threshold value to represent SIH to clarify the effect of SIH on poor prognosis at 30-day,90-day and 1-year.

There was no difference in HbA1c regardless of whether patients had poor outcomes, and this result was also found in another study of acute stroke. This suggests the degree of comorbidities linked to diabetic status rather than HBA1c itself, which associated with poor clinical outcomes^38^. In addition, subgroup analysis demonstrated that the relationship between all indicators and poor clinical outcomes were observed in both diabetic and non-diabetic patients, consistent with other studies^8, 11, 31^. While, many studies currently chose non-diabetic people to analyze SIH to avoid errors caused by the influence of diabetes on blood glucose level, and several studies that included non-diabetic and diabetic people showed inconsistent results in subgroup analyses and revealed that diabetes mellitus affect the relationship^15, 36^. Future interventional studies can be planned to evaluate the effect of diabetes on SIH.

Our experiment innovatively compared the prognostic effect of multiple indicators (including FBG, GG, SHR1 and SHR2) on patients with sICH, and found SHR2(the ratio of FBG to HbA1c) as the most appropriate predictor can be confidently and scientifically selected to represent the association between poor functional outcomes and SIH, which brought a huge breakthrough for future research. However, this study inevitably had some limitations. First, the cohort study based on a Chinese population may restrict its applicability in general. Second, previous studies have indicated that blood glucose fluctuations in patients with ICH may be related to poor prognosis^33^.Because blood glucose was not continuously monitored in this study, dynamic glycemic fluctuation and functional outcomes was not considered. Third, SHR1, SHR2, GG and FBG got different cutoff values at different follow-up times as the threshold for defining SIH, but no further verification was carried out for the obtained threshold values of each indicator in our study.

## Conclusion

This study found that FBG, GG and SHR (both SHR1 and SHR2) were all independently associated with poor functional outcomes in patients with sICH at 30-day,90-day and 1-year.SHR2, the ratio of FBG to HbA1c, could be a trustworthy target for early intervention merit attention in future research, which had a higher predictive value for poor clinical prognosis of sICH at 30 days, 90 days, and 1 year compared with the other three.

## Data Availability

The corresponding authors take full responsibility for the data, the analyses and interpretation, and the conduct of the research. The authors have full access to all of the data and have the right to publish any and all data separate and apart from any sponsor.

## Acknowledgments

We thank all staff members and participants in this study.

